# Metabolic markers distinguish COVID-19 from other intensive care patients and show potential to stratify for disease risk

**DOI:** 10.1101/2021.01.13.21249645

**Authors:** Franziska Schmelter, Bandik Foeh, Alvaro Mallagaray, Johann Rahmoeller, Marc Ehlers, Selina Lehrian, Vera von Kopylow, Inga Künsting, Anne Sophie Lixenfeld, Emily Martin, Mohab Ragab, Max Borsche, Alexander Balck, Eva Juliane Vollstedt, Roza Meyer-Saraei, Fabian Kreutzmann, Ingo Eitel, Stefan Taube, Christine Klein, Alexander Katalinic, Jan Rupp, Eckard Jantzen, Tobias Graf, Christian Sina, Ulrich L Günther

**Author notes:** Institute for Chemistry and Metabolomics, University of Lübeck, Ratzeburger Allee 160, 23562 Lübeck, Germany (Tobias Graf, Christian Sina, Ulrich L Günther). shared first authorship, equal contributions (Franziska Schmelter, Bandik Foeh). made equal contributions (Christian Sina, Ulrich L Günther).

## Abstract

Coronavirus disease 2019 (COVID-19) is a viral infection affecting multiple organ systems of great significance for metabolic processes. Thus. there is increasing interest in metabolic and lipoprotein signatures of the disease and early analyses have demonstrated metabolic pattern typical for atherosclerotic and hepatic damage in COVID-19 patients. However, it remains unclear whether these are specific for COVID-19 or a general marker of critical illness. To answer this question, we have analyzed 276 serum samples from 92 individuals using NMR metabolomics, including longitudinally collected samples from 5 COVID-19 and 11 cardiogenic shock intensive care patients, 18 SARS-CoV-2 antibody-positive individuals, and 58 healthy controls.

COVID-19 patients showed a distinct metabolic serum profile, including changes typical for severe dyslipidemia and a deeply altered metabolic status compared to healthy controls. Specifically, VLDL parameters, IDL particles, large-sized LDL particles, and the ApoB100/ApoA1 ratio were significantly increased, whereas HDL fractions were decreased. Moreover, a similarly perturbed profile was apparent, even when compared to other ICU patients suffering from cardiogenic shock, highlighting the impact of COVID-19 especially on lipid metabolism and energy status. COVID-19 patients were separated with an AUROC of 1.0 when compared to both healthy controls and cardiogenic shock patients. Anti-SARS-CoV-2 antibody-positive individuals without acute COVID-19 did not show a significantly perturbed metabolic profile compared to age- and sex-matched healthy controls, but SARS-CoV-2 antibody-titers correlated significantly with metabolic parameters, including levels of glycine, ApoA2, and small-sized LDL and HDL subfractions. Our data suggest that NMR metabolic profiles are suitable for COVID-19 patient stratification and post-treatment monitoring.

## Introduction

Although SARS-CoV-2 is a primarily respiratory virus, it has become evident that it does not exclusively affect the airways and lungs but a multitude of organs throughout the body^1,2^. Accordingly, severe cases of Coronavirus disease 2019 (COVID-19) are often characterized by multi-organ damage. In the lungs COVID-19 causes acute respiratory distress arising from inflammatory cell infiltration dominated by lymphocytes, diffuse alveolar damage, and pulmonary edema^3^. Moreover, COVID-19 patients show neurological symptoms^4^, renal^5–7^, and liver damage^8^. Vascular damage and thromboembolisms contribute to the observed multi-organ damage and overall lethality of the disease^9^.

This raises the question whether there is a common underlying molecular mechanism that is reflected in metabolic profiles. Early proteomics studies showed dysregulation of coagulative and proinflammatory pathways, associated with metabolic suppression and dyslipidemia linked to the clinical severity of the disease^10,11^. Metabolic profiles from COVID-19 patients were shown to be distinctly different from those of controls, with metabolic markers indicative of liver dysfunction, dyslipidemia, diabetes, and coronary heart disease risk^12^. In a Spanish cohort of hospitalized COVID-19 patients increased ketone bodies and markers of dyslipidemia were reported^13^. Metabolic profiles were characterized by reduced levels of aromatic amino acids and increased levels of glutamic acid, phenylalanine, and glucose. Lipoprotein analyses showed increased VLDL, and IDL (sub)-fractions and decreased HDL-levels in both studies^12,13^. These results underline systemic defects caused by COVID-19 and demonstrate the suitability of metabolomics for detection and potentially stratification of COVID-19 patients.

To date, publicly available metabolomics data is limited to the comparison of COVID-19 and healthy controls. However, it is crucially important to identify whether the associated differences in metabolic profiles are specific for COVID-19 and whether markers can be identified that distinguish COVID-19 from other diseases that affect cardiovascular parameters. It is equally important to identify whether metabolic and lipoprotein markers are consistent for COVID-19 patient samples derived from cohorts in different countries. Moreover, we would like to know how samples from individuals tested positive for COVID-19 without developing a severe form of the disease have a different profile compared to patients requiring intensive care.

Here, we utilize NMR metabolomics to analyze the metabolic and lipoprotein profiles of COVID-19 patients (COVID-19) hospitalized at the intensive care unit (ICU) of the University Hospital of Schleswig-Holstein (Campus Lübeck) compared to a group of healthy controls (HC) and a cohort of cardiogenic shock patients (CS) treated in the same ICU but tested negative for SARS-CoV-2. Importantly, we also compared metabolic profiles from asymptomatic individuals who tested positive for Anti-SARS-CoV-2 antibodies (Anti-S1 IgG+) with antibody-negative individuals. Our results show a severely disturbed serum profile, typical for a perturbed energy status, and dyslipidemia in COVID-19 patients compared to HC and even compared to CS, highlighting the impact of COVID-19 on systemic metabolism. Furthermore, antibody titers of Anti-SARS-CoV-2 antibody-positive individuals correlated with markers of metabolic health.

## Results

### Study population

The study population summarized in **Table 1** included SARS-CoV-2 RNA-negative healthy controls (HC), ICU patients with COVID-19 (COVID-19), and ICU patients with cardiogenic shock (CS). The mean age of COVID-19 patients was 59.2 years and did not differ significantly from HC and CS patients (HC: 50.3 y, p = 0.231; CS: 69.9 y, p = 0.091). All study groups were overweight with average BMIs of 25.2 (HC), 26.1 (CS), and 29.6 (COVID-19) kg/m^2^ and no significant differences between COVID-19 patients and control groups were observed (vs HC: p = 0.060; vs CS: p = 0.204). The Simplified Acute Physiology Score II (SAPS II) and the Sepsis-Related Organ Failure Assessment Score (SOFA) were calculated for COVID-19 and CS patients in the ICU to classify disease severity and no significant differences were found (**Table 1**).

**Table 1:**
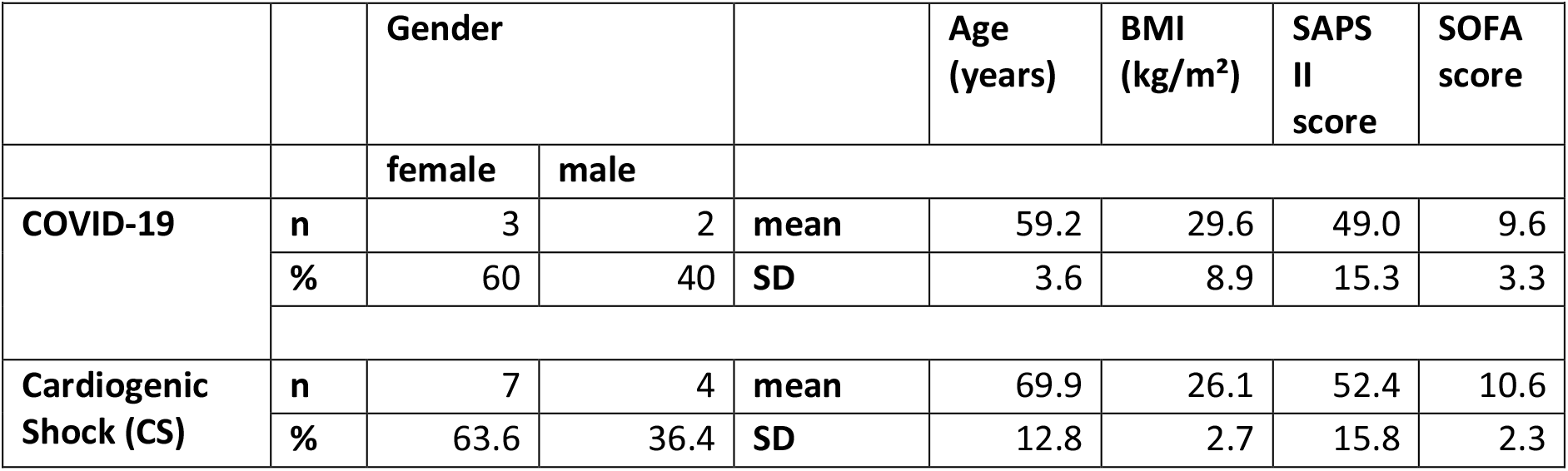
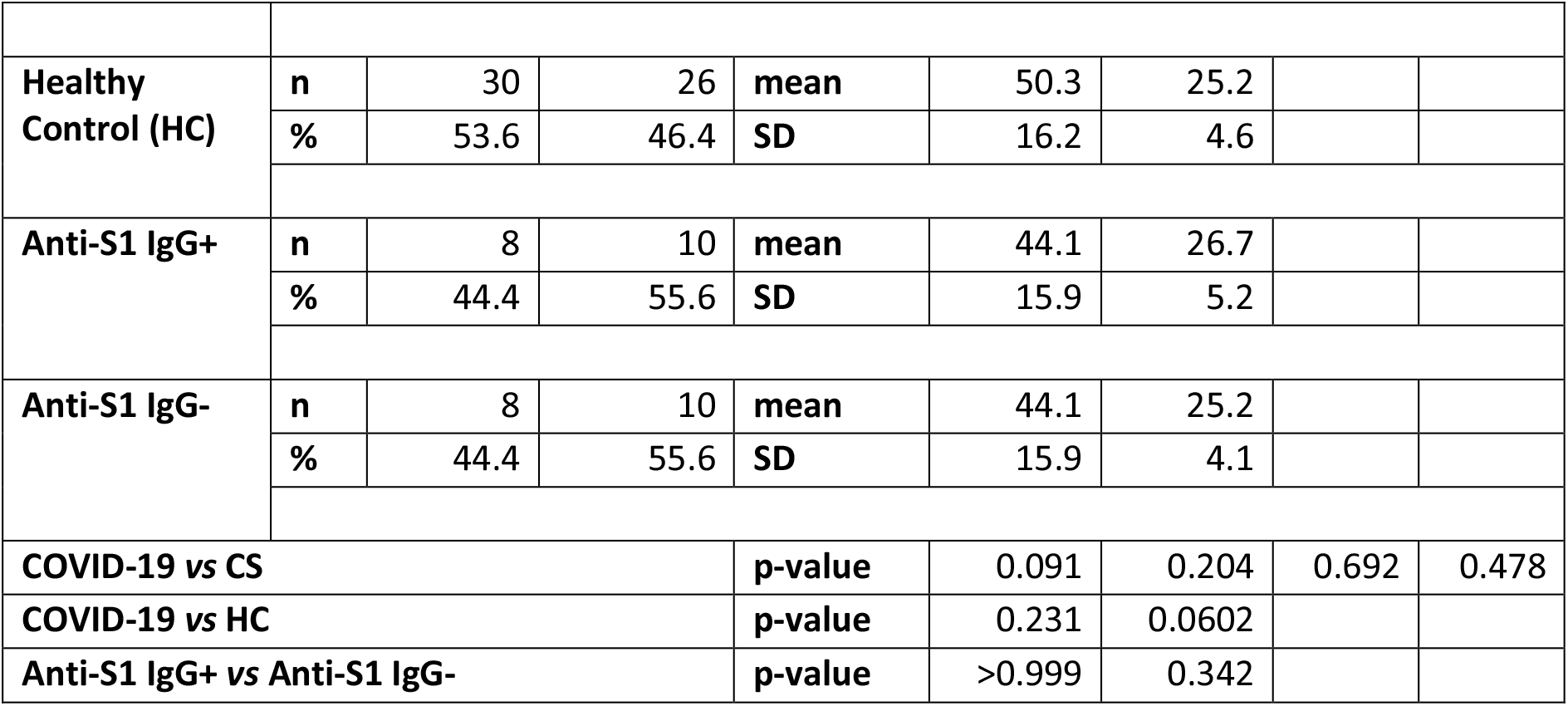
Study population metrics.

| Table 1: Study population metrics |  |  |  |  |  |  |  |  |
| --- | --- | --- | --- | --- | --- | --- | --- | --- |
|  |  | Gender |  |  | Age (years) | BMI (kg/m<sup>2</sup>) | SAPS II score | SOFA score |
|  |  | female | male |  |  |  |  |  |
| COVID-19 | n | 3 | 2 | mean | 59.2 | 29.6 | 49.0 | 9.6 |
|  | % | 60 | 40 | SD | 3.6 | 8.9 | 15.3 | 3.3 |
| Cardiogenic Shock (CS) | n | 7 | 4 | mean | 69.9 | 26.1 | 52.4 | 10.6 |
|  | % | 63.6 | 36.4 | SD | 12.8 | 2.7 | 15.8 | 2.3 |
| Healthy Control (HC) | n | 30 | 26 | mean | 50.3 | 25.2 |  |  |
|  | % | 53.6 | 46.4 | SD | 16.2 | 4.6 |  |  |
| Anti-S1 IgG+ | n | 8 | 10 | mean | 44.1 | 26.7 |  |  |
|  | % | 44.4 | 55.6 | SD | 15.9 | 5.2 |  |  |
| Anti-S1 IgG- | n | 8 | 10 | mean | 44.1 | 25.2 |  |  |
|  | % | 44.4 | 55.6 | SD | 15.9 | 4.1 |  |  |
| COVID-19 vs CS |  |  |  | p-value | 0.091 | 0.204 | 0.692 | 0.478 |
| COVID-19 vs HC |  |  |  | p-value | 0.231 | 0.0602 |  |  |
| Anti-S1 IgG+ vs Anti-S1 IgG- |  |  |  | p-value | >0.999 | 0.342 |  |  |

Additionally, we included asymptomatic individuals that tested positive for Anti-SARS-CoV-2 antibodies (Anti-S1 IgG+) and age- and sex-matched controls (Anti-S1 IgG-) who were both tested negative for SARS-CoV-2 RNA on the same day. Anti-S1-IgG+ individuals had a slightly, but not significantly higher BMI compared to Anti-S1-IgG-(Anti-S1-IgG+: 26.7 kg/m^2^, Anti-S1-IgG-: 25.2 kg/m^2^, p = 0.342).

### Untargeted NMR metabolomics and lipidomics distinguish between COVID-19, HC, and CS

To test whether there are distinct metabolic changes in SARS-CoV-2 infected patients, NMR spectra from blood samples of COVID-19, CS, and HC were acquired. PCA plots of the primary 1D NOESY spectra showed good separation between ICU patients with COVID-19 from HC, but also between COVID-19 and CS patients treated at the ICU during the same period (**Fig. 1A**). Visible differences between the groups were readily observed in 1D NOESY spectra as shown in **Fig. 1B**.

**Figure 1.**
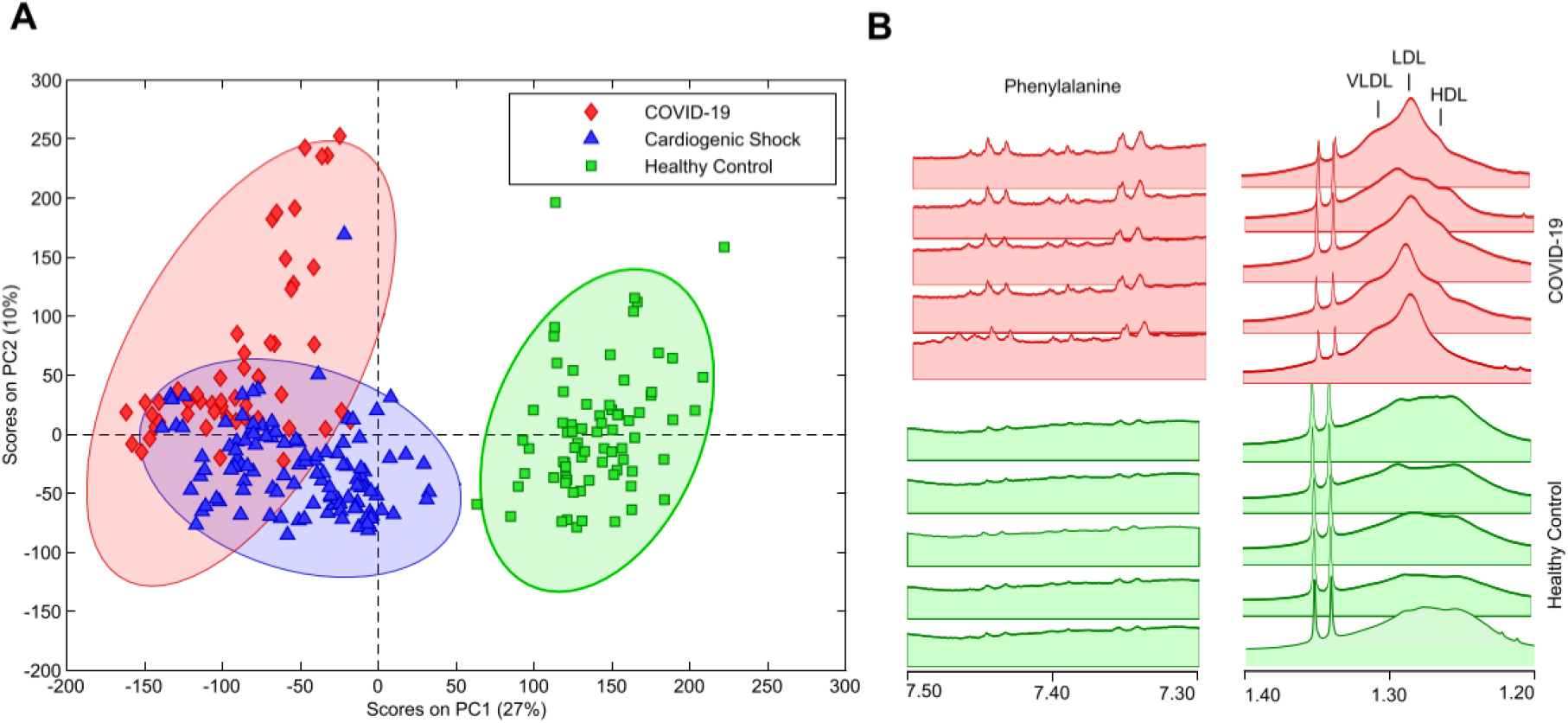
**A**. Untargeted Principal Component Analysis of primary 1D-NOESY NMR spectra for COVID-19 patients (red), Cardiogenic Shock patients (blue), and Healthy Controls (green). **B**. 1D-NOESY spectra showing typical differences between the HC and COVID-19 group.

Principal Component (PC) 1 accounted for 27% of the total variation and discriminated between longitudinally acquired COVID-19 samples and both HC and CS patients, whereas PC2 accounted for 10% of the total variation and predominantly discriminated between COVID-19 and CS patients. These results indicate distinct changes in the metabolic and lipoprotein profiles of COVID-19 patients compared to healthy individuals and similarly severe cases of CS. A similar PCA generated for CPMG spectra (not shown) yields a significantly worse separation suggesting that lipoprotein signals contribute significantly to the PCA classification.

### Targeted analysis reveals metabolite and lipoprotein clusters separating COVID-19 patients from HC and ICU CS patients

To identify metabolites and lipoproteins that are changed in COVID-19 patients, several metabolites and lipoproteins were quantified from NMR spectra. In **Fig. 2A** variance-scaled PCA of the identified and quantified parameters (Table S1) confirmed a clear separation of COVID-19 patients from HC. Notably, the three COVID-19 samples that could not be distinguished from healthy controls by PCA (circled in red) were from one COVID-19 patient with less severe symptoms, who was kept at the ICU only for observation (**Fig. 2A**). Loading plots and hierarchical clustering for PCA revealed several clusters of amino acids, lipoprotein fractions, and subfractions that were most influential for group separation (**Fig. 2B+C**). Moreover, PLS-DA of the full set of metabolites and lipoproteins confirmed the most significant discriminating metabolites. Cross-validation yielded an area under the ROC curve (AUROC) of 1.00 indicative of a near-perfect separation of COVID-19 from HC (**Fig. S1A+B**). To evaluate the specificity of metabolic changes in COVID-19 patients, the metabolic profiles were compared with those from a group of SARS-CoV-2-negative ICU patients suffering from respiratory distress due to CS. Unsupervised PCA of the targeted analysis confirmed good separation in the first two PCs (**Fig. 2D**), in agreement with the separation observed for the PCA on raw data (**Fig. 1A**). Hierarchical clustering revealed amino acid and lipoprotein clusters that separated COVID-19 from CS (**Fig. 2E+F**), in large parts recapitulating the relevant class indicators for separating COVID-19 and HC (**Fig. 2E+F**). PLS-DA confirmed class separation between COVID-19 and CS with an area under the ROC of 0.99, indicative for a distinct separation (**Fig. S1C+D**). In general, the trajectories of repeated measurements in COVID-19 patients moved further away with time as shown representatively for one patient (*Pat. 17*) in **Fig. 2D**, whereas the trajectories of CS patients remained stationary during their stay at the ICU (comp. *Pat. 07*, **Fig. 2D**).

**Figure 2.**
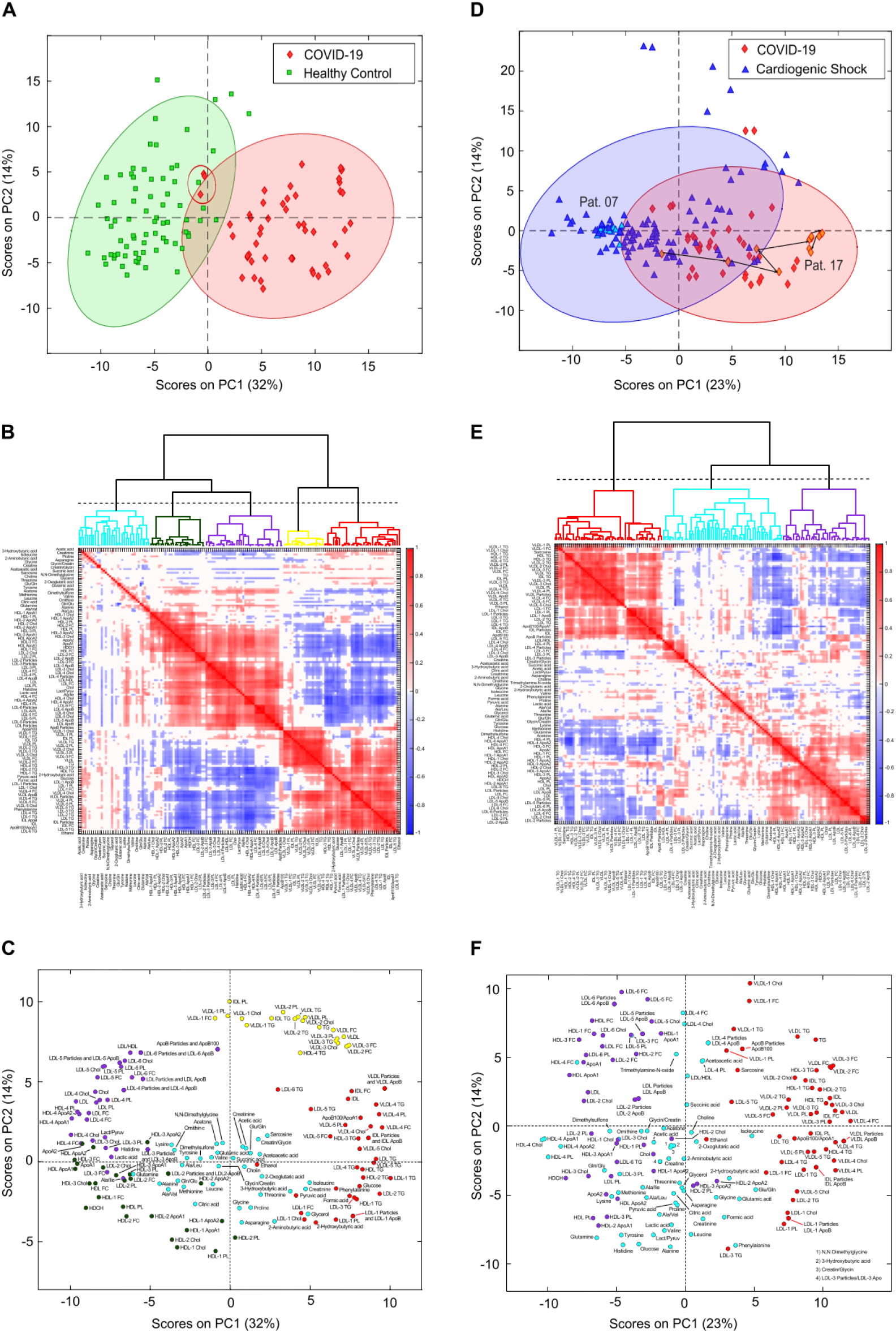
Targeted principal component and cluster analysis for IVDr NMR data. **A**. Scores plot of COVID-19 samples (red) *vs* Healthy Controls (green) showing almost perfect separation of the two groups. **B**. Corresponding hierarchical clustering of correlations for the loadings plot (**C**) of the same PCA showing groups of correlated metabolites and lipoproteins. **D**. PCA scores plot of COVID-19 (red) *vs* Cardiogenic Shock samples (blue) showing partial separation of the two groups. **E**. Corresponding hierarchical clustering of correlations for the loadings plot (**D**) of the same PCA showing groups of correlated metabolites and lipoproteins.

### Disrupted energy status and severe dyslipidemia distinguish COVID-19 patients from HC and ICU CS patients

To examine the metabolic effects of COVID-19 more closely, pairwise analyses of metabolites and lipoproteins were carried out. In **Fig. 3** Forest plots indicate changes between COVID-19 and HC (blue circles) as well as between COVID-19 and CS (red diamonds) (showing relative changes scaled by standard deviation. The reference for COVID-19 *vs* HC is the mean of HC, whereas for COVID-19 *vs* CS the reference line represents the mean of CS). For COVID-19 glucose and formic acid levels were increased and lactic acid and the lactic acid/pyruvic acid ratio decreased compared to HC indicating a disturbed energy status. Decreased glutamine, histidine and Fischer’s ratio ((valine + leucine + isoleucine) / (phenylalanine + tyrosine)) were indicative of disrupted hepatic amino acid metabolism and hepatic damage ^12^. Increased phenylalanine levels and decreased glutamine/glutamate ratios for both HC and CS patients further support a disrupted hepatic metabolism even compared to critically ill CS patients (**Fig. 3A**). Together these results indicate a disturbed energy status and possibly hepatic damage in COVID-19 patients.

**Figure 3.**
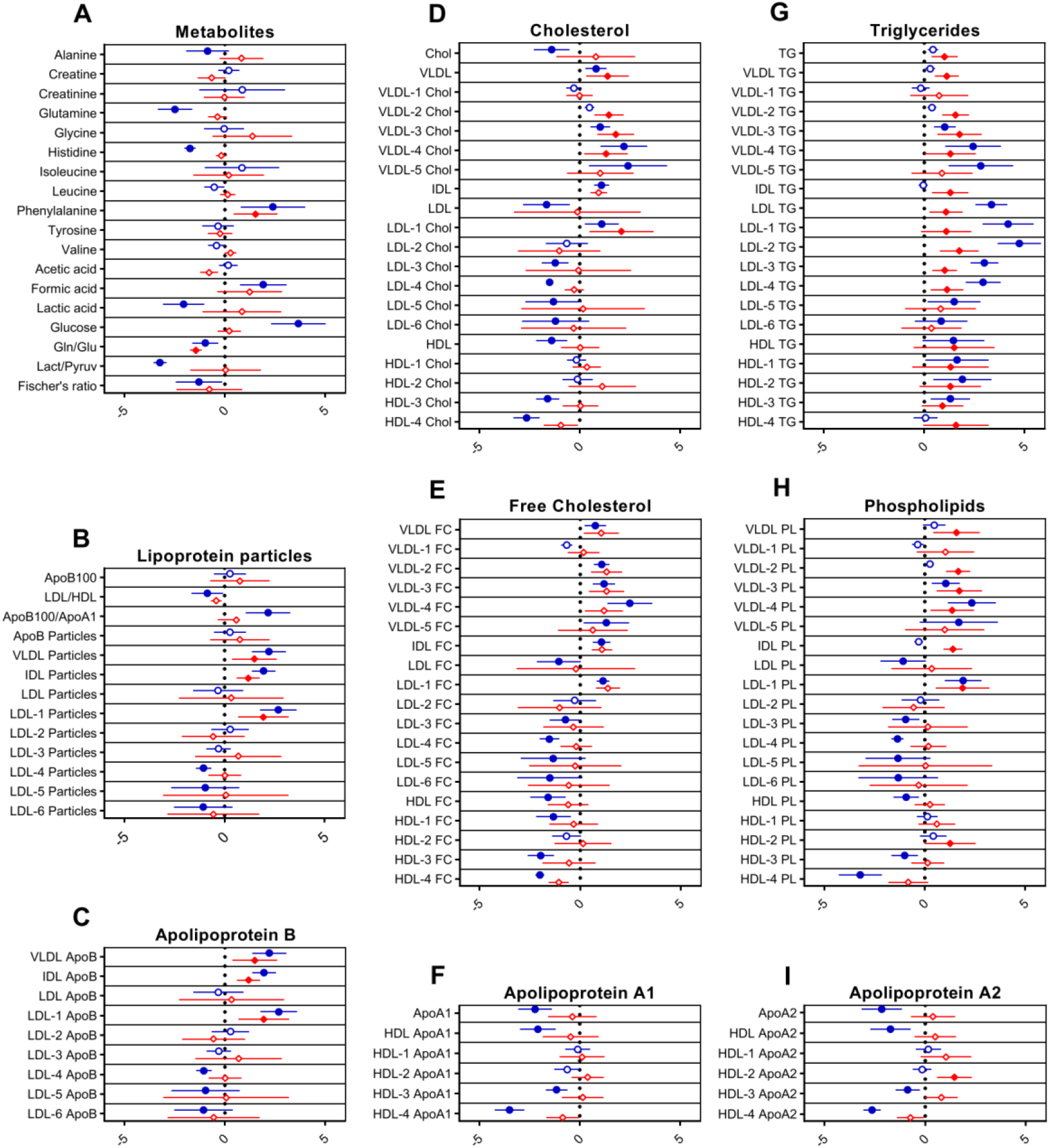
Forest plots showing changes for groups of metabolites (**A**) and lipoprotein classes (**B-I**). The middle line indicates the reference average whereas circles and diamonds on the horizontal axes show changes scaled by standard deviations. Blue circles show changes for COVID-19 samples with Healthy Controls as the reference (vertical line). Red diamonds show changes for COVID-19 with Cardiogenic Shock patients used as the reference (vertical line). Statistically significant differences determined using the false discovery method of Benjamini and Hochberg (Q = 15%) are indicated by filled circles or diamonds.

Lipoprotein profiles of COVID-19 patients showed markers of severe dyslipidemia when compared to HC. VLDL and IDL particle numbers were increased significantly, whereas LDL particle numbers remained unchanged (**Fig. 3B**). The ratio of apolipoprotein B100 to apolipoprotein A1 (ApoB100/ApoA1), a strong independent risk factor for cardiovascular diseases, was significantly increased (**Fig. 3B**). Triglycerides for nearly all lipoprotein subfractions were distinctly elevated, most pronounced for LDL (**Fig. 3G**). HDL, LDL, and several subfractions of HDL and LDL cholesterol (**Fig. 3D+E**), and phospholipids (**Fig. 3H**) were decreased (especially for smaller size particles including LDL-3, 4, 5, and 6, with largest effects observed for HDL-4 cholesterol and HDL-4 phospholipids). Accordingly, levels of apolipoproteins A1 and A2, and B bound in smaller-sized HDL and LDL were reduced (largest effects for HDL-4 ApoA1 and A2) (**Fig. 3C+F+I**). Large-sized LDL-1 particles and bound lipids on the other hand were increased (**Fig. 3B-H**). Subfraction analysis of VLDL lipoproteins further revealed that triglycerides, phospholipids, esterified, and free cholesterol bound to lipoproteins of smaller sizes (VLDL-3, 4, and 5) were increased dramatically (**Fig. 3D, E, G, H**).

Importantly, COVID-19 displayed dyslipidemic changes in comparison to ICU CS patients with similar markers. In **Fig. 3** red diamonds compare COVID-19 against CS samples, i.e. diamonds to the right of the vertical line represent COVID-19 markers showing an increased or additional effect over CS, whereas diamonds left of the reference line show a reduced or additional effect. Generally, we observe enhanced or additional, but never adverse effects in COVID-19 compared to CS.

In particular, IDL and VLDL particles (**Fig. 3B**), smaller-sized VLDL fractions, and VLDL-bound lipids were distinctly increased in COVID-19 (**Fig. 3D, E, G, H**). Moreover, triglyceride levels of nearly all subfractions, large-sized LDL-1 particle numbers, and lipids bound in LDL-1 were further increased. Distinguishing lipoprotein fractions were IDL triglycerides and phospholipids, VLDL-2 phospholipids and cholesterol and HDL-2 ApoA2 which were increased in COVID-19 compared to CS but not to HC.

### Anti-SARS-CoV-2 antibody-titers correlate with metabolomic and lipidomic markers of health in antibody-positive individuals

To test whether COVID-19 might lead to metabolic changes that persist even after asymptomatic infection or mild disease, we analyzed 34 serum samples from 18 Anti-SARS-CoV-2 IgG+ individuals (Anti-S1 IgG+) and compared them to age- and sex-matched controls without Anti-SARS-CoV-2 IgG-antibodies (Anti-S1 IgG-). Both groups were tested PCR-negative for asymptomatic SARS-CoV-2 infection on the same day of blood sampling. PCA showed no separation between the groups and PLS-DA resulted in a model with very poor separation indicating that there are no relevant changes in the overall serum profile of metabolites and lipoproteins (**Fig. S2A-C**).

Possible links between metabolic serum profiles and antibody status were explored by examining associations between antibody titers in Anti-S1 IgG+ samples. Antibody-titers against SARS-CoV-2 correlated inversely with small-sized LDL-6 particles and LDL-6 bound triglycerides, cholesterol, esterified cholesterol, and phospholipids. Similarly, several lipids bound in small-sized HDL-4 were negatively correlated (**Fig. 4**). A strong positive correlation of antibody titers was found for the amino acid glycine, a marker of metabolic health (**Fig. 4P**, p = 0.0062, r = 0.4878). This signature difders from that observed for COVID-19 patients and may suggest an association of metabolic health and Anti-SARS-CoV-2 IgG antibody titers in individuals after asymptomatic infection/mild disease.

**Figure 4.**
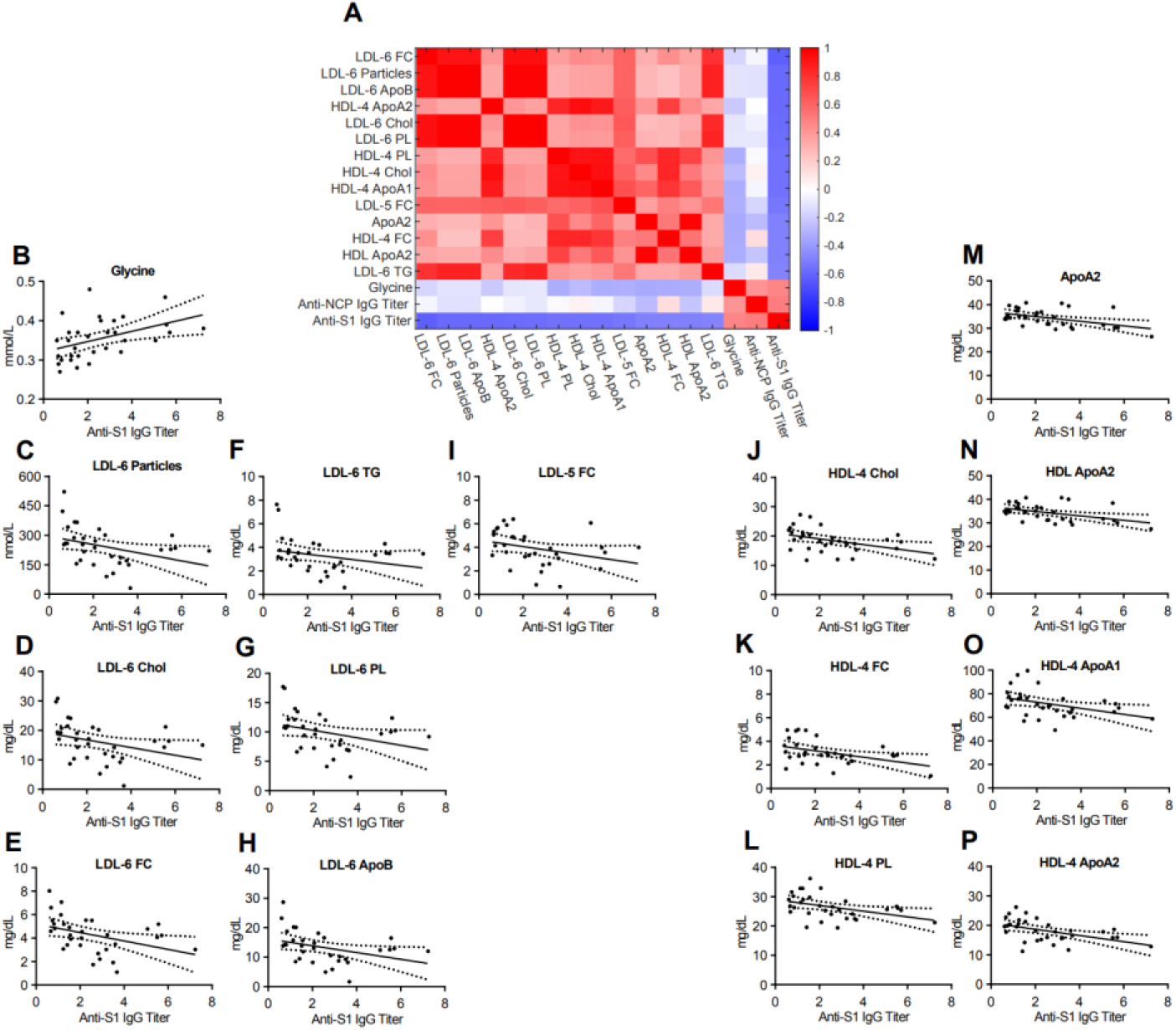
**A**. Heatmap correlating metabolite and lipoprotein levels against the Anti-S1 IgG titer for individuals who tested positive for Anti-SARS-CoV-2 antibodies (Anti-S1 IgG+). **B-P**. Spearman correlation plots for the most significant individual metabolites or lipoproteins against the Anti-S1 IgG titer (p-values in supplementary table).

## Discussion

The emergence of COVID-19 has rapidly developed into a global health crisis affecting health systems to an unprecedented extent. Primarily a respiratory virus, SARS-CoV-2 affects multiple organ systems throughout the body, including the cardiovascular system, the kidney, and the liver. Although these organs are tightly involved and affected by metabolic processes, studies that target metabolism in COVID-19 have been scarce and only looked at differences between COVID-19 patients during infection and healthy controls ^10–13^. Here, we have investigated metabolic changes during and after COVID-19 using ^1^H NMR spectroscopy of serum samples and compared them not only to HC but also to a cohort of critically ill CS patients. The results feature consistent decreases in glutamine/glutamate ratios and increases in phenylalanine levels in COVID-19 across comparisons to both, HC and CS. Furthermore, a deeply disturbed lipoprotein profile typical for dyslipidemia was observed. Finally, antibody titers from mild cases without acute infection correlated with the amino acid glycine and lower levels of small-sized LDL and HDL subclasses.

Early studies based primarily on proteomics data indicated important effects of SARS-CoV-2 infection on the host’s metabolism. On a cellular level SARS-CoV-2 induces changes in protein, carbon, and nucleic acid metabolism in infected host cells enabling rapid viral replication and revealing possible therapeutic targets^14^. *Shen et al*. identified dysregulation of macrophage, platelet degranulation, and complement system pathways, but also observed metabolic suppression indicated by decreased phospholipids, choline, and several amino acids involved in arginine metabolism^10^. Another study found enrichment of metabolites and lipids related to carbon, amino acid, and cholesterol metabolism, as well as hepatic damage^11^, which concurs with clinical features of cardiovascular and hepatic damage frequently observed in COVID-19 patients^4,9,15,16^.

In our data, COVID-19 patients can be clearly separated from both HC and ICU patients with CS using supervised and unsupervised statistical approaches. Targeted analyses revealed a specific profile of COVID-19 patients, that can be distinguished from HC and CS patients. Among other parameters decreased glutamine/glutamate ratios correspond to recently reported results^13^. This ratio is commonly decreased in healthy individuals after physical, anaerobic stress and is associated with an increased risk of losing body cell mass. Low glutamine/glutamate ratios are also typical for a pre-cachectic state in cancer and severe infections^17^. Increased glutamine consumption by activated immune cells can surpass even the rate of glucose consumption and is found in severe infectious disease states^18^. Additionally, the liver switches from a glutamine depot into one of the largest consumers of glutamine in a catabolic disease state^18^, supporting a disturbed, pre-cachectic energy status with a highly active immune system in COVID-19 patients, even compared to CS. It is also possible that the release of alanine-transaminase into the blood stream after liver damage increases deamination of glutamine to form glutamate.

The aromatic amino acid phenylalanine is another parameter specifically increased in COVID-19. It is metabolized to tryptophan almost exclusively in the liver and is elevated in patients with decreased liver function^19^, possibly indicating hepatic damage in the COVID-19 group. Supporting data stems from three independent studies that have shown phenylalanine to be elevated in COVID-19 patients compared to HC as well^12,13,20^. Several other metabolites, that have been shown to be dysregulated in COVID-19 agree with our results, including increased levels of glucose, and reduced levels of histidine, and methionine^13,20^. Increased levels of glucose may be interpreted as a sign of a severely disrupted energy metabolism, possibly even glucose tolerance and prediabetes. Reduced levels of histidine and lactic acid are indicative of metabolic suppression in COVID-19 patients supporting results of previous studies^10,13,20^. However, these changes are not recapitulated when comparing COVID-19 patients to CS controls, challenging the specificity of these metabolic markers for COVID-19. Instead, it might be a general effect of critical illness and catabolic state linked to disturbed energy status.

Dyslipidemia was consistently found in previous studies using proteomics and NMR spectroscopy, indicating disturbed lipoprotein metabolism and increased cardiovascular risk in COVID-19 patients compared to healthy controls^10–13^. Our data show a severely disturbed lipoprotein profile with increased TGs, ApoB100/ApoA ratio, VLDL, IDL, and decreased HDL lipoproteins and subfractions compared to HC. Strikingly, this severely dyslipidemic profile is conserved in large parts in comparison to CS patients. VLDL, IDL, and LDL-1 particle numbers are elevated, VLDL and LDL-1 cholesterol, and phospholipids are increased. Moreover, subfraction analysis shows, that COVID-19 increases predominantly small-sized VLDL-subclasses (VLDL-3, 4, and 5), which are more atherogenic than larger particles^21,22^. Ample evidence suggests IDL and small VLDL remnants that are increased in COVID-19 as an important, independent risk factor for cardiovascular diseases and major cardiac injuries^22–27^.

Similarly, high TG levels contribute to atherosclerosis^28,29^ and are increased irrespectively of lipoprotein classes and subfractions in COVID-19 patients compared to HC but remarkably also compared to CS patients. Elevated TG levels are also linked to the formation of particularly atherogenic, small-sized LDLs^27^, which is not observed in our data. In fact, the COVID-19 patients displayed an imbalance towards larger-sized LDLs, indicated by increased numbers of LDL-1 (*vs* HC and CS) and reduced LDL-4, 5, and 6 (only *vs* HC). Although larger-sized LDL particles are implicated to be less atherogenic than small LDL-particles due to reduced infiltration of vascular intima^27^, these results do not suggest a protective phenotype in the overall dyslipidemic context of COVID-19 patients.

Metabolic and lipoprotein profiles from Anti-SARS-CoV-2 antibody-positive patients without acute infection did not differ significantly from antibody-negative healthy controls. This could point towards the normalization of metabolic processes in the aftermath of COVID-19 infection. However, it is important to consider that the antibody-positive cohort consisted of individuals that were only mildly affected by COVID-19 symptoms several weeks before the serum analyses. Considering this, the observed associations of Anti-S1 and Anti-NCP IgG antibody titers with glycine and lipoprotein fractions in those individuals are even more remarkable. Glycine is known to be reduced in patients with typical metabolic disorders (e.g. obesity, diabetes mellitus type II, and non-alcoholic fatty liver disease)^30^, has been shown to reduce cholesterol and lipid levels in rodents^31^, and correlates inversely with risk of acute myocardial infarction in patients with stable Angina pectoris^32^. Therefore, glycine can potentially be considered a marker of metabolic health (Alves et al., 2019) and is currently tested as a therapeutic agent for metabolic syndrome, atherosclerosis, and even COVID-19 (ClinicalTrials.gov Identifier: NCT04443673). Older studies have shown that glycine is involved in enabling the survival and proliferation of lymphocytes and possibly increases antibody production^33,34^. The inverse correlation of antibody titers with LDL especially of smaller sizes (LDL-3 to 6) is another indicator of increased metabolic health in individuals with high titers against SARS-CoV-2. However, at this point we cannot determine cause and effect from the available data. Possibly, individuals in a healthy metabolic state are more likely to mount an effective immune response to SARS-CoV-2 resulting in higher antibody titers after mild or asymptomatic infection. On the other hand, altered metabolites and lipoproteins could be a consequence of a successful immune reaction to SARS-CoV-2 indicated by higher antibody levels. These findings deserve further investigation as potential markers for individuals who are more likely to mount an adequate immune reaction to SARS-CoV-2 vs those who are at risk of developing more severe disease symptoms.

In conclusion, we show that the metabolic profile of COVID-19 patients is critically disrupted showing typical signs of a pre-cachectic catabolic state, hepatic damage, and severe dyslipidemia even compared to intensive care patients suffering from cardiogenic shock. Curiously, antibody titers after mild infection with SARS-CoV-2 correlate with markers of metabolic health. Taken together, these results underline the severity of changes in amino acid and lipoprotein metabolism and further support the relevance of metabolic disruption by COVID-19. There is now mounting evidence from others and ourselves suggesting that NMR metabolic profiles can be used for early identification and possibly for stratification of COVID-19 patients. Further evidence for the applicability of these data to predict the severity of a COVID-19 infection can be expected after longitudinal data of infected individuals becomes available from ongoing studies and sample collections.

## Methods

### Study Participants

All participants provided written informed consent according to the declaration of Helsinki. ICU patients with either COVID-19 or CS were included within the LUERIC-study at the University Hospital of Schleswig-Holstein (Campus Lübeck) after evaluation and approval by the corresponding ethics committee (LUERIC-MICROBIOME Nr. 19-019 and 19-019 A, University of Luebeck). COVID-19 status was determined by RT-PCR from nasopharyngeal swabs and typical symptoms (fever, cough, dyspnea). Serum samples at the ICU were taken twice daily between 7 and 9 AM and PM every day.

Anti-SARS-CoV-2 antibody-positive individuals and healthy controls did not show any symptoms of COVID-19 and were recruited within the ELISA-study framework as approved by the corresponding ethics committee (ELISA Nr. 20-150, University of Luebeck). Asymptomatic SARS-CoV-2 infection was ruled out by RT-PCR from nasopharyngeal swabs on the same day of serum sampling.

### NMR-Metabolomics

Aliquots of the frozen serum samples were thawed at room temperature for several minutes. Samples were mixed 1/1 (v/v) with 75 mM sodium phosphate buffer (pH 7.4) and shaken manually for 1 minute. 600 µL of mixed samples were transferred into a 5 mm NMR tube. NMR analysis was performed on Bruker 600 MHz Avance III HD spectrometer with a TXI probe. A Bruker SampleJet™ automatic sample changer was used and cooling was set at 6 °C. All experiments were recorded using the Bruker *in vitro* Diagnostic Research (IVDr) protocol. For quality control, the NMR spectrometer was calibrated daily using a strict Standard Operation Procedure as described by Dona et al^35^.

Four different ^1^H NMR experiments were measured per sample at a temperature of 310K. One-dimensional (1D) proton spectra were recorded by using a standard Bruker pulse program (noesygppr1d) with presaturation water suppression during the 4 sec recycle delay. A 1D Carr-Purcell-Meiboom-Gill (CPMG) spin-echo experiment (pulseprogram: cpmgpr1d) was acquired for suppression of proteins and other macromolecular signals. These spectra were acquired with 32 scans and 131,072 data points. Furthermore, 2D J-resolved and diffusion-edited experiments were performed. From these spectra concentrations of 39 metabolites and 112 lipoproteins were obtained automatically using B.I.Quant-PS 2.0.0 and the Bruker IVDr Lipoprotein Subclass Analysis B.I.-LISA™ (Bruker BioSpin).

### Anti-SARS-CoV-2 ELISA

Antibody titers against SARS-CoV-2 in serum samples were determined using commercially available enzyme-linked immunoassay (ELISA)-kits. ELISAs for the spike (S1) and nucleocapsid (NCP) antigens were performed following the manufacturer’s protocols (Anti-SARS-CoV-2-ELISA IgG, EI 2606-9601 G; Anti-SARS-CoV-2-NCP-ELISA IgG, EI 2606-9601-2 G; EUROIMMUN Medizinische Labordiagnostika AG, Germany). Individuals were categorized as positive if they reached a reference value of 0.6 compared to a standard serum. The determination of IgG antibody titers was repeated after a six-week interval for patients, who initially tested positive, combined with additional NMR metabolomics.

### Statistics

Primary NMR data was processed, scaled, and aligned using Matlab (TheMathworks) and MetaboLab^36,37^. Principal Component Analysis (PCA) and Partial Least Square-Discriminant Analysis (PLS-DA) were calculated using PLS-Toolbox (Eigenvector Research) in Matlab. To calculate PCA plots for primary one-dimensional (1D) spectra data were mean-centered and scaled by variance. PCA was further applied to variance-scaled concentrations of metabolites and lipoproteins determined by Bruker’s IVDr software. Similarly, PLS-DA was calculated using PLS-Toolbox using venetian blinds for cross-validation and determine the area under the receiver –operator characteristic (AUROC). Uncorrected two-tailed p-values from t-tests <= 0.01 were selected to exclude uncorrelated metabolites. Clusters were defined via hierarchical agglomerative ward linkage clustering of pairwise correlations. A threshold of 2.5 was selected for the comparison of COVID-19 *vs* HC, and a threshold of 3.5 was used for the comparison of COVID-19 *vs* CS.

Correlation plots were generated using routines written in Matlab. Specifically, pairwise correlations were generated using the *corr* function provided by the Matlab Statistics Toolbox using Spearman type ranked correlations. For pairwise comparisons, seven-day means of the longitudinal measurements were calculated for each ICU patient and compared to the respective controls. Pairwise comparisons were done using multiple, unpaired t-tests adjusting for multiple comparisons using the false discovery rate approach in Prism 8.4.3 (GraphPad Software, LLC). Spearman correlation plots and Forest plots were both generated using Prism 8.4.3 (GraphPad Software, LLC).

## Data Availability

Our ethics does not allow to share the data without signing an agreement. Parties interested can contact the corresponding author to obtain access to this data.

## Authorship Contributions

TG, RMS and IE collected the LUERIC samples; BF, MB, EJV, AB, MB, JR, ME, SL, VvK, IK, AL, EM collected the ELISA samples; JR, ME, SL, VvK, IK, AL, EM, ME performed the antibody analysis; FS performed the NMR experiments; BF, FS, AM, UG performed the statistical data analysis; ME, JR, SL, MR performed and analyzed SARS-CoV-2 AB-Test; CS, TG, UG designed the research; ME, ST, CK, AK and JR designed and carried out the ELISA study; TG, RMS, CS, and IE designed the LUERIC study; BF, FS, CS, EJ, ST, IE and UG wrote the paper.

## Acknowledgements

BF and MR are funded by the DFG, IRTG1911 program; CS is Fresenius Kabi endowed Professor for Nutritional Medicine. CS received research funding from Fresenius Kabi GmbH. FS and EJ are employees of GALAB GmbH, Hamburg. CK is funded by the DFG (Cluster of Excellence 2167 and FOR 2488).

## References

1. Prasad, A. & Prasad, M. Single Virus Targeting Multiple Organs: What We Know and Where We Are Heading? Frontiers in Medicine 7, 370 (2020).

2. Zaim, S., Chong, J. H., Sankaranarayanan, V. & Harky, A. COVID-19 and Multiorgan Response. Current Problems in Cardiology vol. 45 (2020).

3. Xu, Z. et al.. Pathological findings of COVID-19 associated with acute respiratory distress syndrome. The Lancet Respiratory Medicine 8, 420–422 (2020).

4. Mao, L. et al.. Neurologic Manifestations of Hospitalized Patients with Coronavirus Disease 2019 in Wuhan, China. JAMA Neurology 77, 683–690 (2020).

5. Ronco, C., Reis, T. & Husain-Syed, F. Management of acute kidney injury in patients with COVID-19. The Lancet Respiratory Medicine vol. 8 738–742 (2020).

6. Benedetti, C., Waldman, M., Zaza, G., Riella, L. V. & Cravedi, P. COVID-19 and the Kidneys: An Update. Frontiers in Medicine vol. 7 423 (2020).

7. Li, Z. et al.. Caution on Kidney Dysfunctions of COVID-19 Patients. SSRN Electronic Journal 2020.02.08.20021212 (2020) doi:10.1101/2020.02.08.20021212.

8. Xiao, F. et al.. Evidence for Gastrointestinal Infection of SARS-CoV-2. Gastroenterology 158, 1831-1833.e3 (2020).

9. Nishiga, M., Wang, D. W., Han, Y., Lewis, D. B. & Wu, J. C. COVID-19 and cardiovascular disease: from basic mechanisms to clinical perspectives. Nature Reviews Cardiology vol. 17 543–558 (2020).

10. Shen, B. et al.. Proteomic and Metabolomic Characterization of COVID-19 Patient Sera. Cell 182, 59-72.e15 (2020).

11. Wu, D. et al.. Plasma metabolomic and lipidomic alterations associated with COVID-19. National Science Review 7, 1157–1168 (2020).

12. Kimhofer, T. et al.. Integrative Modeling of Quantitative Plasma Lipoprotein, Metabolic, and Amino Acid Data Reveals a Multiorgan Pathological Signature of SARS-CoV-2 Infection. J. Proteome Res. acs.jproteome.0c00519 (2020) doi:10.1021/acs.jproteome.0c00519.

13. Bruzzone, C. et al.. SARS-CoV-2 Infection Dysregulates the Metabolomic and Lipidomic Profiles of Serum. iScience 23, 101645 (2020).

14. Bojkova, D. et al.. Proteomics of SARS-CoV-2-infected host cells reveals therapy targets. Nature 583, 469–472 (2020).

15. Xu, L., Liu, J., Lu, M., Yang, D. & Zheng, X. Liver injury during highly pathogenic human coronavirus infections. Liver International 1–7 (2020) doi:10.1111/liv.14435.

16. Shi, S. et al.. Association of Cardiac Injury with Mortality in Hospitalized Patients with COVID-19 in Wuhan, China. JAMA Cardiology 5, 802–810 (2020).

17. Kinscherf, R. et al.. Low plasma glutamine in combination with high glutamate levels indicate risk for loss of body cell mass in healthy individuals: The effect of N-acetyl-cysteine. Journal of Molecular Medicine 74, 393–400 (1996).

18. Cruzat, V., Rogero, M. M., Keane, K. N., Curi, R. & Newsholme, P. Glutamine: Metabolism and immune function, supplementation and clinical translation. Nutrients vol. 10 (2018).

19. Dejong, C. H. C., Van De Poll, M. C. G., Soeters, P. B., Jalan, R. & Olde Damink, S. W. M. Aromatic amino acid metabolism during liver failure. in Journal of Nutrition vol. 137 1579S–1585S (American Institute of Nutrition, 2007).

20. Thomas, T. et al.. COVID-19 infection alters kynurenine and fatty acid metabolism, correlating with IL-6 levels and renal status. JCI Insight 5, e140327 (2020).

21. Hodis, H. N. et al.. Triglyceride- and cholesterol-rich lipoproteins have a differential effect on mild/moderate and severe lesion progression as assessed by quantitative coronary angiography in a controlled trial of lovastatin. Circulation 90, 42–49 (1994).

22. Carmena, R., Duriez, P. & Fruchart, J. C. Atherogenic lipoprotein particles in atherosclerosis. Circulation vol. 109 (2004).

23. Nordestgaard, B. G. & Tybjærg-Hansen, A. IDL, VLDL, chylomicrons and atherosclerosis. European Journal of Epidemiology 8, 92–98 (1992).

24. Joshi, P. H. et al.. Remnant Lipoprotein Cholesterol and Incident Coronary Heart Disease: The Jackson Heart and Framingham Offspring Cohort Studies. Journal of the American Heart Association 5, (2016).

25. Pastori, D. et al.. Remnant Lipoprotein Cholesterol and Cardiovascular and Cerebrovascular Events in Patients with Non-Alcoholic Fatty Liver Disease. Journal of Clinical Medicine 7, 378 (2018).

26. Saeed, A. et al.. Remnant-Like Particle Cholesterol, Low-Density Lipoprotein Triglycerides, and Incident Cardiovascular Disease. Journal of the American College of Cardiology 72, 156–169 (2018).

27. Borén, J. et al.. Low-density lipoproteins cause atherosclerotic cardiovascular disease: Pathophysiological, genetic, and therapeutic insights: A consensus statement from the European Atherosclerosis Society Consensus Panel. European Heart Journal vol. 41 2313–2330 (2020).

28. Nordestgaard, B. G., Benn, M., Schnohr, P. & Tybjærg-Hansen, A. Nonfasting triglycerides and risk of myocardial infarction, ischemic heart disease, and death in men and women. Journal of the American Medical Association 298, 299–308 (2007).

29. Nordestgaard, B. G. & Varbo, A. Triglycerides and cardiovascular disease. The Lancet vol. 384 626–635 (2014).

30. Alves, A., Bassot, A., Bulteau, A. L., Pirola, L. & Morio, B. Glycine metabolism and its alterations in obesity and metabolic diseases. Nutrients 11, (2019).

31. Park, T., Oh, J. & Lee, K. Dietary taurine or glycine supplementation reduces plasma and liver cholesterol and triglyceride concentrations in rats fed a cholesterol-free diet. Nutrition Research 19, 1777–1789 (1999).

32. Ding, Y. et al.. Plasma glycine and risk of acute myocardial infarction in patients with suspected stable angina pectoris. Journal of the American Heart Association 5, (2016).

33. Li, P., Yin, Y. L., Li, D., Kim, W. S. & Wu, G. Amino acids and immune function. British Journal of Nutrition 98, 237–252 (2007).

34. Duval, D., Demangel, C., Munier-Jolain, K., Miossec, S. & Geahel, I. Factors controlling cell proliferation and antibody production in mouse hybridoma cells: I. Influence of the amino acid supply. Biotechnology and Bioengineering 38, 561–570 (1991).

35. Dona, A. C. et al.. Precision high-throughput proton NMR spectroscopy of human urine, serum, and plasma for large-scale metabolic phenotyping. Analytical Chemistry 86, 9887–9894 (2014).

36. Guenther, U. L., Ludwig, C. & Rueterjans, H. NMRLAB - Advanced NMR Data Processing in Matlab. Journal of Magnetic Resonance (2000) doi:10.1006/jmre.2000.2071.

37. Ludwig, C. & Guenther, U. L. MetaboLab - advanced NMR data processing and analysis for metabolomics. BMC Bioinformatics (2011) doi:10.1186/1471-2105-12-366.

