## Supplementary Table and Figures for "Metabolic markers distinguish COVID-19 from other intensive care patients and show potential to stratify for disease risk"

**Table S1.** Descriptive statistics for metabolites and lipoproteins in COVID-19, HC, and CS cohorts**.** For each cohort, mean and standard deviation are shown. P-Values show unadjusted p-values from unpaired t-tests.

| | **COVID-19 (mean)** | SD | **HC (mean)** | SD | **CS (mean)** | SD | **Change in % (vs HC)** | P value (vs HC) | **Change in % (vs CS)** | P value (vs CS) |
|---|---|---|---|---|---|---|---|---|---|---|
| **Metabolites** | | | | | | | | | | |
| Alanine (mmol/l) | 0.45 | 0.11 | 0.53 | 0.10 | 0.35 | 0.13 | -15.91 | *0.07634* | 28.68 | 0.14665 |
| Creatine (mmol/l) | 0.04 | 0.03 | 0.03 | 0.05 | 0.07 | 0.05 | 33.18 | 0.66377 | -43.13 | 0.21856 |
| Creatinine (mmol/l) | 0.11 | 0.04 | 0.09 | 0.02 | 0.11 | 0.06 | 20.13 | 0.10297 | -1.17 | 0.96532 |
| Glutamine (mmol/l) | 0.51 | 0.07 | 0.72 | 0.09 | 0.58 | 0.21 | -29.50 | *0.00000* | -12.46 | 0.46792 |
| Glycine (mmol/l) | 0.36 | 0.08 | 0.36 | 0.08 | 0.29 | 0.05 | -0.82 | 0.94003 | 25.09 | 0.05731 |
| Histidine (mmol/l) | 0.06 | 0.01 | 0.09 | 0.02 | 0.07 | 0.03 | -33.51 | *0.00034* | -6.53 | 0.71834 |
| Isoleucine (mmol/l) | 0.09 | 0.04 | 0.07 | 0.02 | 0.09 | 0.03 | 28.17 | 0.09687 | 7.02 | 0.76120 |
| Leucine (mmol/l) | 0.12 | 0.02 | 0.14 | 0.04 | 0.11 | 0.06 | -13.88 | 0.25804 | 7.95 | 0.76631 |
| Phenylalanine (mmol/l) | 0.13 | 0.03 | 0.08 | 0.02 | 0.08 | 0.03 | 49.70 | *0.00001* | 63.04 | *0.01330* |
| Tyrosine (mmol/l) | 0.06 | 0.01 | 0.07 | 0.02 | 0.07 | 0.03 | -8.36 | 0.47091 | -9.30 | 0.64493 |
| Valine (mmol/l) | 0.26 | 0.02 | 0.28 | 0.06 | 0.23 | 0.11 | -8.03 | 0.37660 | 12.52 | 0.56466 |
| Acetic acid (mmol/l) | 0.01 | 0.01 | 0.01 | 0.02 | 0.03 | 0.02 | 32.17 | 0.70442 | -58.50 | 0.13194 |
| Formic acid (mmol/l) | 0.04 | 0.02 | 0.01 | 0.01 | 0.02 | 0.01 | 231.37 | *0.00016* | 67.36 | *0.06039* |
| Lactic acid (mmol/l) | 2.24 | 0.88 | 3.98 | 0.86 | 1.76 | 0.58 | -43.88 | *0.00006* | 26.86 | 0.21717 |
| Glucose (mmol/l) | 8.90 | 1.66 | 4.43 | 1.23 | 8.10 | 3.69 | 100.87 | *<0.000001* | 9.78 | 0.65771 |
| Glutamine/Glutamate ratio | 3.51 | 1.19 | 5.31 | 1.86 | 10.71 | 5.23 | -33.94 | 0.03855 | -67.22 | *0.00977* |
| Lactic acid/Pyruvic Acid ratio | 22.13 | 6.74 | 89.09 | 20.91 | 21.95 | 5.00 | -75.16 | *<0.000001* | 0.83 | 0.95229 |
| Fischer's ratio | 2.58 | 0.65 | 3.31 | 0.57 | 2.96 | 0.52 | -22.03 | *0.00888* | -12.89 | 0.22863 |
| **Lipoproteins** | | | | | | | | | | |
| ApoB100 (mg/dl) | 91.70 | 19.96 | 84.79 | 25.22 | 78.97 | 17.45 | 8.16 | 0.55406 | 16.12 | 0.21558 |
| LDL/HDL | 1.23 | 0.65 | 1.93 | 0.83 | 2.48 | 3.17 | -36.47 | *0.07038* | -50.36 | 0.40579 |
| ApoB100/ApoA1 | 1.00 | 0.23 | 0.56 | 0.21 | 0.82 | 0.32 | 79.37 | *0.00002* | 21.77 | 0.28030 |
| ApoB Particles (mmol/l) | 1667.36 | 362.85 | 1541.66 | 458.51 | 1435.91 | 317.20 | 8.15 | 0.55413 | 16.12 | 0.21563 |

| | | | | | | | | | | |
|---|---|---|---|---|---|---|---|---|---|---|
| VLDL Particles (mmol/l) | 327.91 | 67.48 | 151.64 | 80.13 | 216.38 | 78.49 | 116.25 | *0.00001* | 51.54 | *0.01600* |
| IDL Particles (mmol/l) | 199.77 | 30.13 | 100.36 | 51.28 | 123.74 | 67.36 | 99.06 | *0.00008* | 61.44 | *0.03192* |
| LDL Particles (mmol/l) | 1160.29 | 487.50 | 1286.14 | 393.10 | 1082.97 | 242.44 | -9.78 | 0.50314 | 7.14 | 0.67197 |
| LDL-1 Particles (mmol/l) | 396.20 | 65.35 | 204.67 | 71.96 | 271.76 | 67.24 | 93.58 | *<0.000001* | 45.79 | *0.00384* |
| LDL-2 Particles (mmol/l) | 198.61 | 60.17 | 180.63 | 64.79 | 225.24 | 49.90 | 9.95 | 0.55265 | -11.82 | 0.36767 |
| LDL-3 Particles (mmol/l) | 157.27 | 44.27 | 179.00 | 73.61 | 139.69 | 26.75 | -12.14 | 0.52047 | 12.59 | 0.33605 |
| LDL-4 Particles (mmol/l) | 72.72 | 40.96 | 183.93 | 107.31 | 71.88 | 65.10 | -60.46 | *0.02577* | 1.17 | 0.97944 |
| LDL-5 Particles (mmol/l) | 120.16 | 171.55 | 217.13 | 101.81 | 115.51 | 71.33 | -44.66 | *0.05916* | 4.02 | 0.93856 |
| LDL-6 Particles (mmol/l) | 183.25 | 208.46 | 334.64 | 143.97 | 246.09 | 118.24 | -45.24 | *0.03377* | -25.53 | 0.44930 |
| Chol (mg/dl) | 142.67 | 40.21 | 205.37 | 45.49 | 122.31 | 26.62 | -30.53 | *0.00424* | 16.64 | 0.24529 |
| VLDL (mg/dl) | 32.46 | 7.19 | 21.32 | 13.81 | 20.78 | 8.82 | 52.29 | *0.08126* | 56.23 | *0.02172* |
| VLDL-1 Chol (mg/dl) | 6.64 | 2.49 | 8.54 | 6.59 | 6.66 | 4.95 | -22.35 | 0.52497 | -0.39 | 0.99149 |
| VLDL-2 Chol (mg/dl) | 4.61 | 0.64 | 3.28 | 2.72 | 3.02 | 1.15 | 40.51 | 0.28310 | 52.78 | *0.01227* |
| VLDL-3 Chol (mg/dl) | 6.29 | 1.35 | 3.45 | 2.79 | 2.99 | 1.93 | 82.31 | *0.02888* | 110.22 | *0.00407* |
| VLDL-4 Chol (mg/dl) | 10.27 | 3.22 | 4.18 | 2.79 | 5.45 | 3.86 | 145.49 | *0.00002* | 88.35 | *0.02952* |
| VLDL-5 Chol (mg/dl) | 2.82 | 1.20 | 1.33 | 0.62 | 1.89 | 0.95 | 112.45 | *0.00002* | 49.04 | 0.11582 |
| IDL (mg/dl) | 21.17 | 2.99 | 12.46 | 8.05 | 12.68 | 9.35 | 69.87 | *0.02023* | 66.91 | *0.07137* |
| LDL (mg/dl) | 53.17 | 40.26 | 110.19 | 34.92 | 54.96 | 16.57 | -51.75 | *0.00101* | -3.26 | 0.89886 |
| LDL-1 Chol (mg/dl) | 29.61 | 6.36 | 21.29 | 7.61 | 19.30 | 5.19 | 39.11 | *0.02116* | 53.44 | *0.00394* |
| LDL-2 Chol (mg/dl) | 13.13 | 7.69 | 17.82 | 7.40 | 17.85 | 4.88 | -26.29 | 0.18129 | -26.45 | 0.15488 |
| LDL-3 Chol (mg/dl) | 7.40 | 5.07 | 16.65 | 7.65 | 7.54 | 2.52 | -55.55 | *0.01058* | -1.91 | 0.93920 |
| LDL-4 Chol (mg/dl) | 1.85 | 1.55 | 15.88 | 9.46 | 2.92 | 4.20 | -88.32 | *0.00171* | -36.58 | 0.59482 |
| LDL-5 Chol (mg/dl) | 6.32 | 11.32 | 16.84 | 8.17 | 5.55 | 4.79 | -62.50 | *0.00961* | 13.82 | 0.84794 |
| LDL-6 Chol (mg/dl) | 10.99 | 14.63 | 21.49 | 8.84 | 13.07 | 7.29 | -48.87 | *0.01918* | -15.94 | 0.70372 |
| HDL (mg/dl) | 39.07 | 12.31 | 61.03 | 15.92 | 38.55 | 16.74 | -35.99 | *0.00399* | 1.34 | 0.95177 |
| HDL-1 Chol (mg/dl) | 19.05 | 4.82 | 20.63 | 10.20 | 16.03 | 8.85 | -7.67 | 0.73400 | 18.80 | 0.49163 |
| HDL-2 Chol (mg/dl) | 9.88 | 2.50 | 10.24 | 3.34 | 7.79 | 1.95 | -3.47 | 0.81779 | 26.78 | 0.08935 |
| HDL-3 Chol (mg/dl) | 6.60 | 1.60 | 11.01 | 2.79 | 6.51 | 2.36 | -40.01 | *0.00102* | 1.51 | 0.93429 |
| HDL-4 Chol (mg/dl) | 3.31 | 3.55 | 17.61 | 5.46 | 8.13 | 5.41 | -81.20 | *<0.000001* | -59.25 | 0.09289 |
| VLDL FC (mg/dl) | 13.26 | 2.75 | 9.33 | 5.23 | 9.13 | 4.11 | 42.13 | 0.10380 | 45.25 | 0.06164 |

| | | | | | | | | | | |
|---|---|---|---|---|---|---|---|---|---|---|
| VLDL-1 FC (mg/dl) | 1.64 | 0.77 | 3.51 | 2.83 | 1.44 | 1.27 | -53.16 | 0.14953 | 14.22 | 0.74694 |
| VLDL-2 FC (mg/dl) | 2.78 | 0.48 | 1.46 | 1.24 | 1.77 | 0.80 | 90.53 | *0.02264* | 56.82 | *0.02237* |
| VLDL-3 FC (mg/dl) | 3.12 | 0.71 | 1.59 | 1.30 | 1.80 | 1.06 | 95.86 | *0.01242* | 73.47 | *0.02431* |
| VLDL-4 FC (mg/dl) | 5.19 | 1.50 | 1.86 | 1.36 | 2.85 | 2.07 | 178.67 | *0.00000* | 82.16 | *0.04095* |
| VLDL-5 FC (mg/dl) | 1.09 | 0.46 | 0.56 | 0.41 | 0.88 | 0.35 | 94.57 | *0.00825* | 23.74 | 0.32774 |
| IDL FC (mg/dl) | 6.22 | 1.02 | 3.73 | 2.35 | 3.50 | 2.63 | 66.78 | *0.02315* | 77.84 | *0.04463* |
| LDL FC (mg/dl) | 21.77 | 10.20 | 31.63 | 9.41 | 22.68 | 4.51 | -31.15 | *0.02957* | -3.98 | 0.80500 |
| LDL-1 FC (mg/dl) | 8.71 | 0.71 | 6.18 | 2.24 | 6.69 | 1.53 | 40.88 | *0.01522* | 30.24 | *0.01484* |
| LDL-2 FC (mg/dl) | 4.70 | 2.50 | 5.33 | 2.36 | 6.22 | 1.56 | -11.86 | 0.56886 | -24.43 | 0.15564 |
| LDL-3 FC (mg/dl) | 3.54 | 1.63 | 5.06 | 2.08 | 3.99 | 1.41 | -30.05 | 0.11845 | -11.31 | 0.57929 |
| LDL-4 FC (mg/dl) | 1.27 | 1.07 | 4.64 | 2.23 | 1.61 | 1.78 | -72.61 | *0.00151* | -20.82 | 0.70622 |
| LDL-5 FC (mg/dl) | 1.88 | 3.09 | 4.46 | 1.94 | 2.29 | 1.76 | -57.79 | *0.00873* | -17.88 | 0.73726 |
| LDL-6 FC (mg/dl) | 2.41 | 3.22 | 5.41 | 2.01 | 3.52 | 2.06 | -55.48 | *0.00349* | -31.57 | 0.41451 |
| HDL FC (mg/dl) | 7.07 | 3.95 | 14.18 | 4.48 | 9.95 | 5.07 | -50.14 | *0.00113* | -28.97 | 0.28150 |
| HDL-1 FC (mg/dl) | 2.35 | 2.07 | 5.52 | 2.41 | 3.04 | 2.22 | -57.38 | *0.00610* | -22.54 | 0.56986 |
| HDL-2 FC (mg/dl) | 1.71 | 0.65 | 2.33 | 0.92 | 1.63 | 0.60 | -26.35 | 0.14972 | 4.99 | 0.80915 |
| HDL-3 FC (mg/dl) | 0.60 | 0.55 | 2.26 | 0.85 | 0.89 | 0.55 | -73.50 | *0.00008* | -32.54 | 0.34498 |
| HDL-4 FC (mg/dl) | 0.26 | 0.30 | 2.93 | 1.35 | 1.05 | 0.77 | -91.19 | *0.00005* | -75.34 | *0.04777* |
| TG (mg/dl) | 187.08 | 22.26 | 148.36 | 89.21 | 143.95 | 44.56 | 26.10 | 0.34046 | 29.96 | *0.06239* |
| VLDL TG (mg/dl) | 110.42 | 15.24 | 91.92 | 62.32 | 74.81 | 33.28 | 20.13 | 0.51355 | 47.60 | *0.04068* |
| VLDL-1 TG (mg/dl) | 44.26 | 17.52 | 51.15 | 40.02 | 32.96 | 15.82 | -13.46 | 0.70598 | 34.29 | 0.22013 |
| VLDL-2 TG (mg/dl) | 18.54 | 2.49 | 14.09 | 11.64 | 11.28 | 4.88 | 31.62 | 0.39982 | 64.40 | *0.00769* |
| VLDL-3 TG (mg/dl) | 20.44 | 4.74 | 11.45 | 8.82 | 11.07 | 5.58 | 78.50 | *0.02899* | 84.63 | *0.00588* |
| VLDL-4 TG (mg/dl) | 19.32 | 6.31 | 8.20 | 4.60 | 11.28 | 6.47 | 135.49 | *0.00001* | 71.29 | *0.03601* |
| VLDL-5 TG (mg/dl) | 5.21 | 1.42 | 2.72 | 0.89 | 4.18 | 1.22 | 91.84 | *<0.000001* | 24.80 | 0.15667 |
| IDL TG (mg/dl) | 16.21 | 3.47 | 16.97 | 15.95 | 10.05 | 4.97 | -4.48 | 0.91631 | 61.32 | *0.02615* |
| LDL TG (mg/dl) | 48.23 | 6.63 | 20.28 | 8.42 | 37.42 | 10.46 | 137.82 | *<0.000001* | 28.90 | *0.05374* |
| LDL-1 TG (mg/dl) | 17.98 | 3.77 | 5.58 | 2.99 | 13.88 | 3.93 | 222.18 | *<0.000001* | 29.51 | *0.07080* |
| LDL-2 TG (mg/dl) | 7.16 | 1.04 | 2.62 | 0.97 | 4.83 | 1.40 | 173.05 | *<0.000001* | 48.42 | *0.00516* |
| LDL-3 TG (mg/dl) | 5.25 | 0.62 | 2.57 | 0.90 | 3.99 | 1.30 | 104.52 | *<0.000001* | 31.67 | *0.06077* |

| | | | | | | | | | | |
|---|---|---|---|---|---|---|---|---|---|---|
| LDL-4 TG (mg/dl) | 6.96 | 1.31 | 2.54 | 1.52 | 4.69 | 2.10 | 173.73 | *<0.000001* | 48.38 | *0.04454* |
| LDL-5 TG (mg/dl) | 5.10 | 2.05 | 2.79 | 1.57 | 3.93 | 1.51 | 83.21 | *0.00304* | 29.84 | 0.21678 |
| LDL-6 TG (mg/dl) | 5.95 | 2.51 | 4.33 | 1.93 | 5.20 | 2.18 | 37.41 | 0.08359 | 14.49 | 0.54907 |
| HDL TG (mg/dl) | 22.01 | 9.21 | 13.30 | 6.04 | 13.54 | 5.96 | 65.40 | *0.00448* | 62.55 | *0.04271* |
| HDL-1 TG (mg/dl) | 9.56 | 4.56 | 4.82 | 2.92 | 5.65 | 3.13 | 98.43 | *0.00156* | 69.22 | *0.06337* |
| HDL-2 TG (mg/dl) | 4.73 | 1.73 | 2.47 | 1.21 | 2.92 | 1.47 | 91.90 | *0.00026* | 62.29 | *0.04717* |
| HDL-3 TG (mg/dl) | 4.01 | 1.10 | 2.55 | 1.13 | 2.81 | 1.38 | 57.24 | *0.00759* | 42.88 | 0.10971 |
| HDL-4 TG (mg/dl) | 3.65 | 0.82 | 3.56 | 1.37 | 2.66 | 0.65 | 2.66 | 0.88046 | 37.05 | *0.02086* |
| VLDL PL (mg/dl) | 27.09 | 6.39 | 21.69 | 11.50 | 16.26 | 7.17 | 24.90 | 0.30698 | 66.54 | *0.01201* |
| VLDL-1 PL (mg/dl) | 5.93 | 1.78 | 8.10 | 6.33 | 4.32 | 1.62 | -26.88 | 0.44947 | 37.01 | 0.09730 |
| VLDL-2 PL (mg/dl) | 4.34 | 0.54 | 3.65 | 2.69 | 2.50 | 1.16 | 18.99 | 0.57046 | 73.98 | *0.00478* |
| VLDL-3 PL (mg/dl) | 6.56 | 1.72 | 3.99 | 2.47 | 3.30 | 1.98 | 64.52 | *0.02709* | 98.73 | *0.00685* |
| VLDL-4 PL (mg/dl) | 9.87 | 2.78 | 4.43 | 2.34 | 5.53 | 3.32 | 122.59 | *0.00001* | 78.59 | *0.02379* |
| VLDL-5 PL (mg/dl) | 2.84 | 1.34 | 1.67 | 0.69 | 2.00 | 0.88 | 69.48 | *0.00162* | 41.97 | 0.15469 |
| IDL PL (mg/dl) | 7.23 | 1.01 | 8.62 | 4.68 | 3.48 | 2.79 | -16.13 | 0.51352 | 107.52 | *0.01239* |
| LDL PL (mg/dl) | 44.83 | 19.36 | 63.13 | 17.27 | 40.66 | 12.54 | -28.99 | *0.02817* | 10.25 | 0.60999 |
| LDL-1 PL (mg/dl) | 19.95 | 3.62 | 12.37 | 3.99 | 13.59 | 3.53 | 61.27 | *0.00013* | 46.80 | *0.00509* |
| LDL-2 PL (mg/dl) | 9.36 | 3.40 | 10.13 | 3.65 | 10.86 | 2.85 | -7.55 | 0.65419 | -13.80 | 0.37318 |
| LDL-3 PL (mg/dl) | 5.98 | 2.56 | 9.50 | 3.78 | 5.73 | 1.68 | -37.08 | *0.04633* | 4.35 | 0.81814 |
| LDL-4 PL (mg/dl) | 2.62 | 1.45 | 9.04 | 4.80 | 2.23 | 2.12 | -71.07 | *0.00438* | 17.07 | 0.72233 |
| LDL-5 PL (mg/dl) | 4.10 | 6.60 | 9.53 | 4.13 | 4.00 | 2.59 | -57.03 | *0.00951* | 2.48 | 0.96537 |
| LDL-6 PL (mg/dl) | 7.01 | 8.55 | 12.71 | 4.35 | 8.29 | 4.56 | -44.85 | *0.01278* | -15.48 | 0.69647 |
| HDL PL (mg/dl) | 68.43 | 13.47 | 87.85 | 21.38 | 62.74 | 23.24 | -22.11 | *0.05151* | 9.07 | 0.62175 |
| HDL-1 PL (mg/dl) | 28.15 | 6.84 | 26.47 | 13.28 | 22.52 | 9.73 | 6.35 | 0.78174 | 25.04 | 0.26478 |
| HDL-2 PL (mg/dl) | 18.54 | 3.45 | 16.34 | 5.24 | 14.28 | 3.53 | 13.47 | 0.36309 | 29.85 | *0.04081* |
| HDL-3 PL (mg/dl) | 13.61 | 2.89 | 17.98 | 4.43 | 12.95 | 4.58 | -24.33 | *0.03495* | 5.04 | 0.77555 |
| HDL-4 PL (mg/dl) | 8.20 | 6.04 | 26.21 | 5.67 | 14.39 | 7.92 | -68.71 | *<0.000001* | -42.99 | 0.14500 |
| ApoA1 (mg/dl) | 95.60 | 22.96 | 156.65 | 27.77 | 104.11 | 24.71 | -38.97 | *0.00001* | -8.18 | 0.52521 |
| HDL ApoA1 (mg/dl) | 94.73 | 26.72 | 157.20 | 30.36 | 105.39 | 24.97 | -39.74 | *0.00004* | -10.12 | 0.45085 |
| HDL-1 ApoA1 (mg/dl) | 32.72 | 12.00 | 34.58 | 19.31 | 31.13 | 13.99 | -5.38 | 0.83365 | 5.11 | 0.82961 |

| | | | | | | | | | |
|---|---|---|---|---|---|---|---|---|---|
| HDL-2 ApoA1 (mg/dl) | 18.65 | 3.94 | 22.43 | 6.16 | 16.21 | 6.38 | -16.84 | 0.18481 | 15.03 | 0.44811 |
| HDL-3 ApoA1 (mg/dl) | 21.47 | 3.75 | 29.39 | 6.97 | 20.77 | 4.71 | -26.96 | *0.01541* | 3.36 | 0.77567 |
| HDL-4 ApoA1 (mg/dl) | 20.15 | 10.45 | 69.56 | 14.27 | 33.55 | 16.63 | -71.03 | *<0.000001* | -39.93 | 0.12281 |
| ApoA2 (mg/dl) | 20.96 | 6.17 | 34.23 | 6.22 | 18.22 | 7.33 | -38.76 | *0.00003* | 15.06 | 0.48049 |
| HDL ApoA2 (mg/dl) | 24.20 | 5.94 | 34.36 | 5.97 | 20.61 | 7.54 | -29.56 | *0.00056* | 17.43 | 0.36527 |
| HDL-1 ApoA2 (mg/dl) | 4.10 | 1.25 | 3.77 | 2.00 | 2.82 | 1.29 | 8.66 | 0.72196 | 45.17 | 0.08589 |
| HDL-2 ApoA2 (mg/dl) | 4.26 | 0.66 | 4.47 | 1.41 | 2.88 | 0.99 | -4.75 | 0.74167 | 47.76 | *0.01378* |
| HDL-3 ApoA2 (mg/dl) | 5.83 | 1.06 | 7.35 | 1.77 | 4.52 | 1.69 | -20.74 | *0.06444* | 28.94 | 0.13742 |
| HDL-4 ApoA2 (mg/dl) | 4.83 | 2.10 | 17.72 | 4.94 | 7.66 | 4.04 | -72.77 | *<0.000001* | -36.97 | 0.16638 |
| VLDL ApoB (mg/dl) | 18.03 | 3.71 | 8.34 | 4.41 | 11.90 | 4.32 | 116.25 | *0.00001* | 51.54 | *0.01599* |
| IDL ApoB (mg/dl) | 10.99 | 1.66 | 5.52 | 2.82 | 6.81 | 3.70 | 99.05 | *0.00008* | 61.45 | *0.03190* |
| LDL ApoB (mg/dl) | 63.81 | 26.81 | 70.73 | 21.62 | 59.56 | 13.33 | -9.78 | 0.50323 | 7.14 | 0.67192 |
| LDL-1 ApoB (mg/dl) | 21.79 | 3.59 | 11.26 | 3.96 | 14.95 | 3.70 | 93.58 | *<0.000001* | 45.79 | *0.00384* |
| LDL-2 ApoB (mg/dl) | 10.92 | 3.31 | 9.93 | 3.56 | 12.39 | 2.75 | 9.96 | 0.55225 | -11.82 | 0.36790 |
| LDL-3 ApoB (mg/dl) | 8.65 | 2.43 | 9.84 | 4.05 | 7.68 | 1.47 | -12.13 | 0.52065 | 12.58 | 0.33609 |
| LDL-4 ApoB (mg/dl) | 4.00 | 2.25 | 10.12 | 5.90 | 3.95 | 3.58 | -60.47 | *0.02577* | 1.15 | 0.97977 |
| LDL-5 ApoB (mg/dl) | 6.61 | 9.43 | 11.94 | 5.60 | 6.35 | 3.92 | -44.67 | *0.05914* | 4.01 | 0.93868 |
| LDL-6 ApoB (mg/dl) | 10.08 | 11.46 | 18.40 | 7.92 | 13.53 | 6.50 | -45.24 | *0.03376* | -25.53 | 0.44932 |

**Table S2.** Spearman Correlation for metabolites and lipoproteins against the Anti-S1 IgG-titer in COVID-19, HC, and CS cohorts.

| | Glycine | ApoA2 | LDL-6 Particles | HDL-ApoA2 | LDL-6 TG | LDL-6 Chol | LDL-5 FC | LDL-6 FC | LDL-6 PL | LDL-6 ApoB | HDL-4 Chol | HDL-4 FC | HDL-4 PL | ApoA1 | ApoA2 |
|---|---|---|---|---|---|---|---|---|---|---|---|---|---|---|---|
| Spearman r | 0.5101 | -0.4146 | -0.5099 | -0.4126 | -0.3905 | -0.5051 | -0.4593 | -0.5535 | -0.4938 | -0.5099 | -0.4949 | -0.4092 | -0.4952 | -0.5011 | -0.5257 |
| P value | | | | | | | | | | | | | | | |
| P (two-tailed) | 0.0029 | 0.0183 | 0.0029 | 0.0189 | 0.0271 | 0.0032 | 0.0082 | 0.001 | 0.0041 | 0.0029 | 0.004 | 0.0201 | 0.004 | 0.0035 | 0.002 |

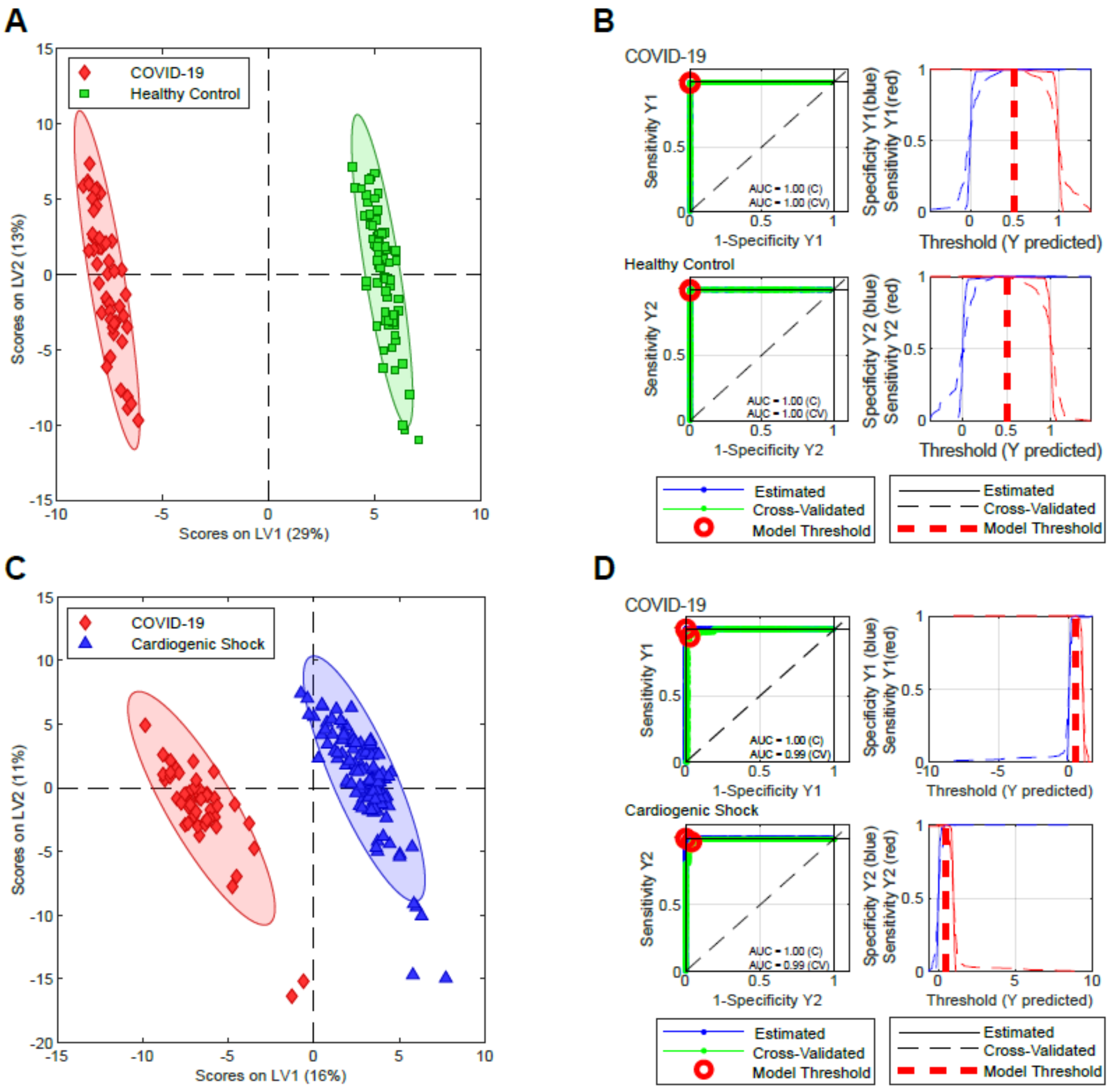

**Supplementary Figure 1. A.** PLS-DA analysis of COVID-19 vs Healthy Control samples and cross-validation derived ROC curves (**B**) showing an area under the ROC curve of 1.00 indicating a perfect separation between groups. **C** and **D**: PLS-DA and ROC analysis for COVID-19 vs Cardiogenic Shock samples.

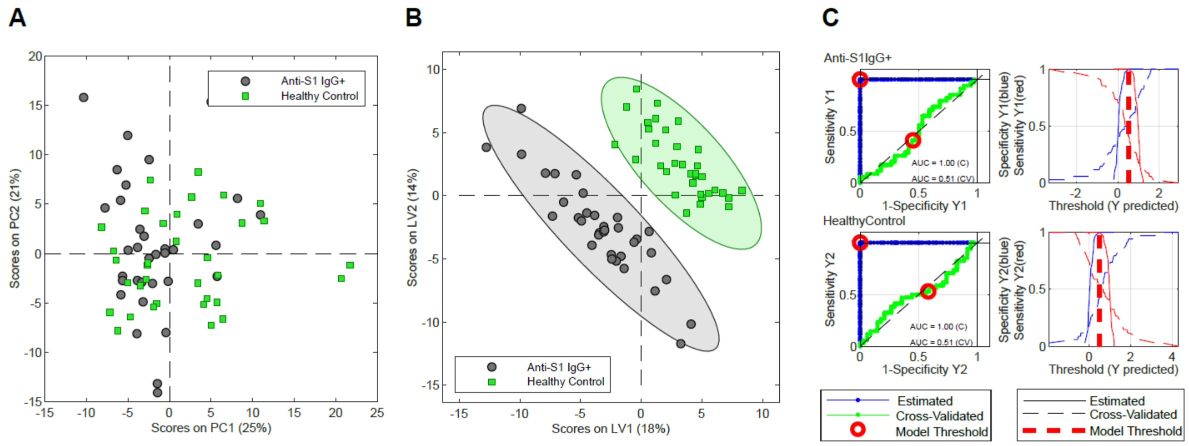

**Supplementary Figure 2. A** PCA and **B** PLS-DA for Anti-S1 IgG+ vs age- and sex-matched Healthy Controls. The ROC analysis in **C** shows no significance for the separation of the two groups.
